# Sustained dengue transmission and seroprevalence in the U.S. Virgin Islands

**DOI:** 10.64898/2026.02.07.26345802

**Authors:** Valerie V. Mac, Joshua M. Wong, Emma S. Jones, Brad J. Biggerstaff, Mark Delorey, Matt D.T. Hitchings, Zachary J. Madewell, Janice Perez-Padilla, Hannah R. Volkman, Freddy A. Medina, Jorge Munoz-Jordan, Brian Wakeman, Valentine Wanga, Aubrey Drummond, Joy Joseph, Annellie Gumbs, Esther M. Ellis, Laura E. Adams

## Abstract

**Objective:** To estimate dengue virus (DENV) seroprevalence and assess parental vaccine perceptions among children in the US Virgin Islands (USVI).

**Methods:** A cross-sectional serosurvey was conducted during April–May 2022 among 372 children aged 8–16 years from 15 schools across USVI. Past DENV infection was determined using a dengue IgG rapid diagnostic test. Data on demographics, dengue knowledge, and vaccine acceptance were collected from parents. Catalytic models estimated annual DENV force of infection (FOI) using seroprevalence and case data from 2010–2022.

**Results:** Seroprevalence among children aged 8–13 years was 47% (95% CI: 29%, 68%). FOI peaked during 2012–2013 outbreaks and remained low in other years. Only 17% of parents were aware of an approved dengue vaccine, and 25% reported they would vaccinate their child.

**Conclusions:** Nearly half of children had prior DENV infection. Despite this risk, parental awareness of dengue vaccination was low, underscoring gaps in detection, diagnosis, and preparedness for vaccine implementation.

**Policy Implications:** These data highlight the need for enhanced surveillance, public education, and targeted planning for dengue vaccine introduction in endemic US jurisdictions.

## INTRODUCTION

Dengue, caused by four antigenically distinct but closely related dengue viruses (DENV-1 to DENV-4) transmitted by *Aedes* species mosquitoes, is the most common arthropod-borne virus (arbovirus) globally.^1,2^ Transmission is sustained in tropical and sub-tropical regions with competent vector populations. Individuals can be infected multiple times during their lifetime, as infection with one DENV serotype confers long-lived protection against that serotype but only transient, partial protection against others. Dengue remains a major global health challenge: the global incidence of dengue doubled from 1990 to 2021, primarily in tropical and subtropical regions.^3^ Although most infections are asymptomatic or mild, about 25% present with a nonspecific febrile illness, and approximately 5% progress to severe dengue, characterized by plasma leakage, shock, hemorrhage, or severe organ involvement.^4,5^

Dengue is endemic in the U.S. Virgin Islands (USVI), a U.S. territory of about 87,000 people,^6,7^ with two major population centers: St. Croix and the combined district of St. Thomas and St. John islands. An outbreak in 2005 led to 331 suspected cases, with about half requiring hospitalization, and one death in a 14-year-old female with severe manifestations due to secondary DENV infection.^8^ From 2010 to 2020, 353 dengue cases were reported from USVI to CDC,^9^ with incidence peaking during the 2012–2013 outbreak. Children and adolescents were disproportionately affected, with 21.5% of cases in the 10–19-year-old age group, although this group comprised only 11.4% of the USVI population in the 2020 Census.^10^ A school-based survey during this period found that 20% of students (40/203) and 17% of the staff (20/118) had anti-DENV IgM, suggesting recent infections within the preceding three months.^11^

Despite past outbreaks and an outbreak declared in August 2024—with 216 locally acquired cases reported that year and 47 additional cases in 2025^12^—no formal, population-based dengue serosurveys have been conducted in USVI. Seroprevalence data are important for public health and healthcare planning and for anticipating resource needs and costs.^13,14^ Seroprevalence estimates can guide targeted vector management activities in high-transmission areas, inform public health education strategies, and help identify populations at greatest risk for secondary infections, thereby supporting decisions about vaccine implementation and anticipating needs for diagnostic testing and clinical care during outbreaks.

Seroprevalence data are particularly critical for planning dengue vaccination strategies, a growing priority in dengue prevention. Dengvaxia, the only currently licensed dengue vaccine in the U.S., is recommended by the Advisory Committee on Immunization Practices (ACIP) for individuals aged 9–16 years living in endemic areas who have laboratory-confirmed prior DENV infection.^15^ The population-level impact of this vaccine depends not only on accurate prevaccination screening but also on sufficiently high seroprevalence among the target population to ensure safety and effectiveness.^16^ Although Dengvaxia manufacturing is being discontinued,^17^ several other dengue vaccines are in development or undergoing licensure, including Qdenga, which is recommended by WHO for routine use in children aged 6–16 years in high-transmission settings but has shown variable efficacy by prior infection history.^18,19,20^ Understanding local seroprevalence is therefore essential to inform decisions about introducing new vaccines and achieving equitable, high-impact implementation.

We conducted a serosurvey to estimate prior DENV infection among school-aged children in the USVI—a population of particular interest for dengue vaccination, as current vaccine strategies focus on children and adolescents. In addition to estimating seroprevalence, we used catalytic modeling to assess the annual force of infection (FOI) and identify risk factors associated with prior infection. We also assessed parental awareness of dengue and evaluated vaccine acceptability to inform future immunization strategies.

## METHODS

### Setting and Community Engagement

During April–May 2022, we conducted a school-based dengue serosurvey in USVI among children in 3^rd^–7^th^ grades across the two health districts: St. Croix Island and St. Thomas/St. John Islands. Ten schools were selected using a single-stage cluster sampling design stratified by the two health districts, with inclusion probabilities proportional to 3rd grade enrollment size. An additional five schools were selected by the Virgin Islands Department of Health (VIDOH) to ensure geographic and demographic representation. Final design weights for the 15 selected schools were derived from 10,000 simulations of the sampling inclusion process and were subsequently adjusted by raking to the age and sex distributions of the 2022 US Census population estimates for the two districts.^6,21^

All selected schools were offered the Dengue Day program—an interactive science education initiative designed to teach students about DENV, the role of epidemiologists, mosquito biology, source reduction, and bite prevention. Participation in Dengue Day was open to all 3^rd^-7^th^ grade students at selected schools, regardless of participation in the serosurvey. This educational approach aimed to equip students with practical knowledge about dengue prevention and empower them to take action in their communities. Informational materials were also sent home to parents, including guidance on dengue symptoms and warning signs of severe illness. At each school, parents received a permission form detailing the serosurvey procedures and a questionnaire to collect demographic information and assess knowledge and perceptions regarding dengue and dengue vaccination. As part of the recruitment process, parents were offered an online question-and-answer session with study staff to learn more about the serosurvey and address any questions before deciding on participation. Participants received a $10 gift card to compensate for the time required to complete the questionnaire. The VIDOH notified parents of their child’s test results.

### Laboratory testing

Children with signed parental permission forms were tested for prior DENV infection using the CTK Biotech OnSite Dengue IgG Rapid Test, which has a sensitivity of 89.6% and specificity of 95.7%.^22^ Testing was performed using 5μL of whole blood obtained via fingerstick. Blood sample collection and test administration occurred onsite at the selected schools. Seropositivity was defined as a positive IgG result, verified independently by 2 trained readers. Discordant readings were adjudicated by the on-site laboratory lead, who assigned the final result.

### Analysis

*Univariate analysis and weighted seroprevalence*: We performed univariate analyses to describe self-reported participant characteristics by health district (St. Croix and St. Thomas/St. John) and for USVI overall. Characteristics included age, sex, race, ethnicity, school type, time living in USVI, and birthplace. To estimate age-stratified seroprevalence of DENV infection among children in grades 3–7, we calculated weighted seroprevalence using the participant IgG test results. To provide an interpretive benchmark relevant to Dengvaxia prevaccination screening, we used the test-performance targets described in CDC’s ACIP dengue vaccine recommendations (PPV ≥90%, with sensitivity ≥75% and specificity ≥98%).^15^ Under these performance targets, a PPV of 90% is achieved when underlying seroprevalence is approximately 20%; therefore, we used 20% seroprevalence as a reference threshold when interpreting our seroprevalence estimates.

*Surveillance case data for catalytic model inputs*: Dengue has been a nationally notifiable disease in the United States since 2010. State and territorial health departments report cases to CDC through the National Arbovirus Surveillance System (ArboNET). Dengue cases were classified according to the Council of State and Territorial Epidemiologists’ (CSTE) 2015 case definition,^23^ meeting specified clinical and laboratory criteria.^23,24^ For the catalytic model, we included locally-acquired cases reported to ArboNET from USVI between 2010 and 2022.

*Catalytic model to estimate force of infection and catalytic model regression analysis:* We estimated the annual dengue FOI using catalytic models informed by individual-level seropositivity data and age-stratified dengue case data from 2010–2022. The annual FOI is equivalent to the hazard function, quantifying the proportion of uninfected (seronegative) individuals who become infected in a given year. Our approach followed methods described by Adams et al. and Kada et al.^13,25^ adapted to account for residence history, diagnostic test performance, and survey design. Briefly, we estimated FOI by modeling cumulative dengue hazard over a child’s lifetime, stratified by health district and prior residence in endemic vs. non-endemic countries, and using the child’s serostatus to infer their lifetime hazard. Bayesian Markov chain Monte Carlo (MCMC) methods were used for parameter estimation, implemented in RStan.^26^ All analyses were performed in R software version 4.2.2 (R Foundation for Statistical Computing, Vienna, Austria). Additional methodological details are provided in Supplemental Methods.

*Vaccine Acceptance and Dengue Perceptions*: Parents were asked four questions related to dengue knowledge and vaccine attitudes: (1) whether they had ever heard of dengue, (2) whether their child had ever had dengue, (3) whether they knew an approved dengue vaccine was recommended for children in the U.S., and (4) whether they would have their child vaccinated if the approved dengue vaccine were made available in the USVI. For vaccine acceptance, parents selected one of three responses: “Yes”, “Unsure”, or “No.” We assessed whether the distribution of vaccine acceptance responses differed by individual demographic characteristics; due to limited sample size, multivariable models were not fit. We used vector generalized linear models to regress vaccine acceptance (Yes, Unsure, No) on each demographic variable, using “Unsure” as the reference category.^27,28^ Likelihood ratio tests compared univariate models against the null; odds ratios were then estimated for each significant predictor. Odds ratios comparing Yes:Unsure and No:Unsure were estimated across levels of each variable. The method described by Zou and Donner^29^ was applied to design-based estimates and standard errors to derive 95% confidence bounds for the odds ratios. Response distributions between levels of a variable were considered statistically different if at least one of the odds ratios was statistically different from 1 at the 95% confidence interval; these estimates have not been adjusted for multiple comparisons. A binomial model was used to examine vaccine acceptance by race categories to account for 0 parents of Asian children responding “undecided”.

## RESULTS

### Participant characteristics and estimated seroprevalence

During April–May 2022, the Dengue Day program reached 1,391 students across USVI, and 372 children aged 8–13 years were enrolled in the serosurvey—192 from St. Croix and 180 from St. Thomas/St. John (Table 1). Most participants (69%) were enrolled in public schools. The sample had slightly more females (55%) and had the highest proportion of participants (27%) among children 10 years old. The majority of participants were Black/African American (75%), followed by White (13%), and other/multiple race groups. Nearly one in four identified as Hispanic or Latino (24%). Most participants were born in USVI (72%) and most had lived there more than 5 years (83%). Among those born outside the USVI, 46% had resided locally for over five years.

**Table 1.** Descriptive characteristics of serosurvey participants by health district — U.S. Virgin Islands, April–May 2022 (N=372).

|  | **Total**<br>**Sample**<br>**n (%)** | **St. Croix**<br>**n (%)** | **St. Thomas/**<br>**St. John**<br>**n (%)** |
| --- | --- | --- | --- |
| **Total** | 372 (100) | 192 (100) | 180 (100) |
| **Age, years** |  |  |  |
| 8 | 56 (15) | 24 (12) | 32 (18) |
| 9 | 76 (20) | 37 (19) | 39 (22) |
| 10 | 100 (27) | 57 (30) | 43 (24) |
| 11 | 52 (14) | 32 (17) | 20 (11) |
| 12 | 58 (16) | 27 (14) | 31 (17) |
| 13 | 30 (8) | 15 (8) | 15 (8) |
| **Sex** |  |  |  |
| Female | 204 (55) | 109 (57) | 95 (53) |
| Male | 168 (45) | 83 (43) | 85 (47) |
| **Race** |  |  |  |
| Black/African-American | 279 (75) | 142 (74) | 137 (76) |
| White | 48 (13) | 24 (12) | 24 (13) |
| Asian/Pacific Islander | 8 (2) | 4 (2) | 4 (2) |
| American Indian/Alaska Native | 0 (0) | 0 (0) | 0 (0) |
| Multiple | 9 (2) | 6 (3) | 3 (2) |
| Other | 28 (8) | 16 (8) | 12 (7) |
| **Ethnicity** |  |  |  |
| Hispanic or Latino | 89 (24) | 55 (29) | 34 (19) |
| Non-Hispanic or Latino | 206 (55) | 102 (53) | 104 (58) |
| Unknown | 77 (21) | 35 (18) | 42 (23) |
| **School Type** |  |  |  |
| Public | 255 (69) | 115 (60) | 140 (78) |
| Private | 117 (31) | 77 (40) | 40 (22) |
| **Birthplace** |  |  |  |
| US | 54 (15) | 32 (17) | 22 (12) |
| USVI | 269 (72) | 149 (78) | 120 (67) |
| Other | 49 (13) | 11 (6) | 38 (21) |
| **Time Living in USVI (All participants)** |  |  |  |
| <1 year | 11 (3) | 6 (3) | 5 (3) |
| 1-5 years | 49 (13) | 18 (9) | 31 (18) |
| >5 years | 309 (83) | 168 (88) | 141 (80) |
| Not disclosed | 3 (1) | --- | --- |
| **Time Living in USVI (participants born outside of USVI)<sup>a</sup> (n=99)** |  |  |  |
| <1 year | 9 (9) | 5 (12) | 4 (7) |
| 1-5 years | 45 (46) | 16 (39) | 28 (49) |
| >5 years | 45 (46) | 20 (49) | 25 (44) |

<sup>a</sup> Among participants not born in the USVI (including those born in U.S. states/territories outside the USVI and those born outside the U.S.); time living in the USVI was self-reported and did not distinguish continuous from cumulative residence

Detection of anti-DENV IgG antibodies revealed substantial prior dengue exposure among participants. The overall weighted seroprevalence among children aged 8–13 years was 47% (95% CI: 29%, 68%) (Figure 1, Supplemental Table A). Among children aged 9–13 years, the age group within this population eligible for Dengvaxia, the estimated seroprevalence was slightly higher at 51% (95% CI: 39%, 64%). Seroprevalence increased with age, ranging from a low of 27% (95% CI: 17%, 39%) among 8-year-olds to a high of 69% (95% CI: 45%, 88%) among 12-year-olds. Estimated seroprevalence was 34% (95% CI: 21%, 50%) in St. Croix and 59% (95% CI: 30%, 80%) in St. Thomas/St. John for all participants. Among female participants, seroprevalence was 50% (95% CI: 22%, 80%); among male participants, seroprevalence was 45% (95% CI: 31%, 59%) (Table SA).

**Figure 1.**
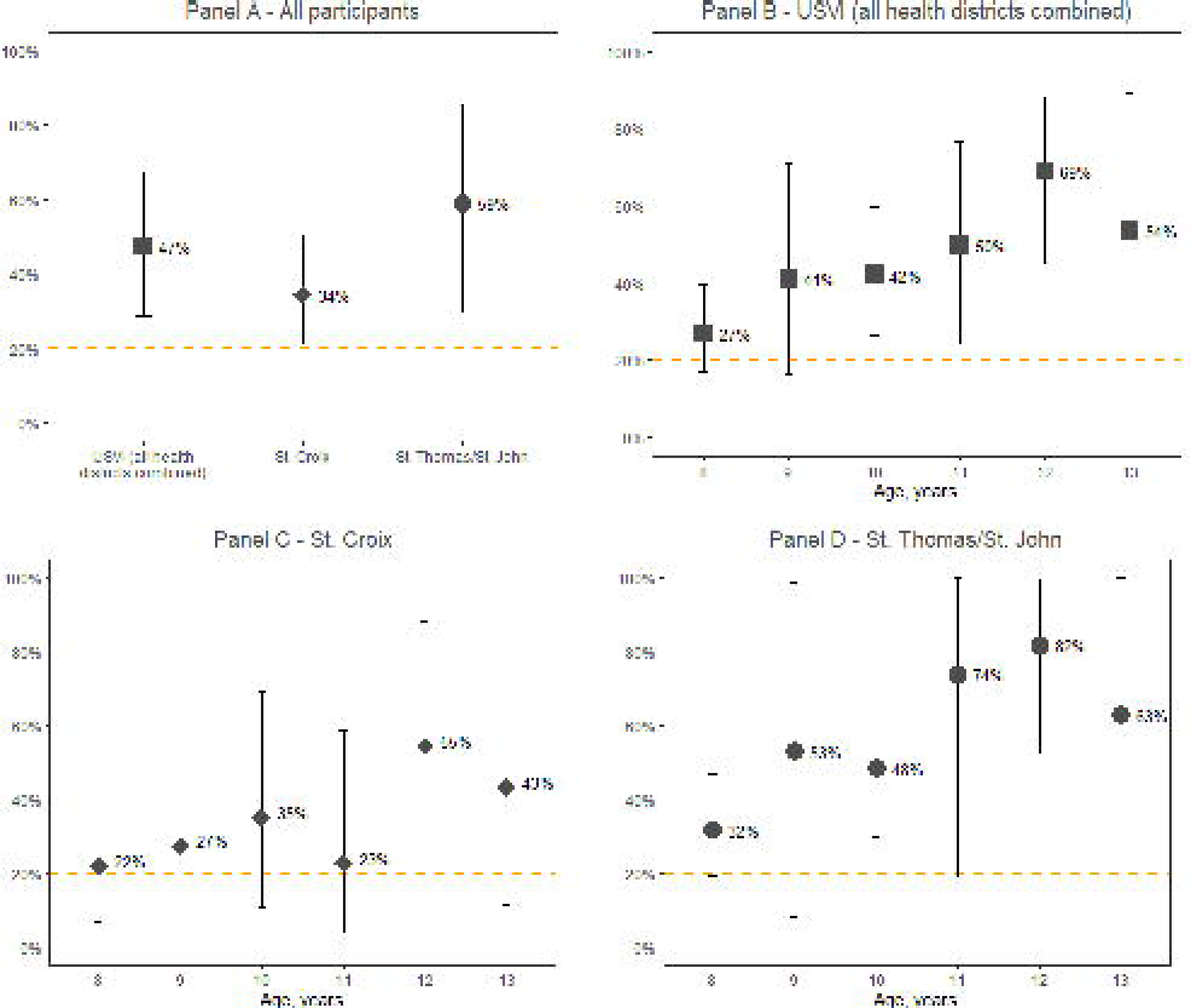
DENV IgG seroprevalence estimates among participants aged 8–13 years overall, in the USVI, and by health district and age group — U.S. Virgin Islands, April–May 2022 (n=372). Seroprevalence estimates are weighted and standardized to the 2022 U.S. Census age and sex distribution across the two health districts, and adjusted for test performance (CTK Biotech OnSite Dengue IgG Rapid Test: sensitivity 89.6%, specificity 95.7%). The dotted line at 20% indicates the minimum seroprevalence at which a prevaccination screening test meeting the ACIP-recommended minimum performance (≥75% sensitivity, ≥98% specificity) would achieve a positive predictive value (PPV) ≥90%, meaning that at least 9 of 10 screen-positive children truly have prior DENV infection. **Panel A:** Seroprevalence estimates for all participants aged 8–13 years (n=372) and stratified by health district (St. Croix [n=192], St. Thomas/St. John [n=180]). **Panel B:** Seroprevalence estimates by single year of age, all districts combined. **Panel C:** Seroprevalence estimates by single year of age, St. Croix only. **Panel D:** Seroprevalence estimates by single year of age, St. Thomas/St. John only.

### Annual dengue transmission and factors associated with risk

Seroprevalence data from this study and case data from passive surveillance showed a consistent pattern, indicating generally limited dengue transmission across USVI outside known outbreak periods (Figure 2, Supplemental Table B). The highest FOI occurred during the 2012–2013 dengue outbreaks, with estimates of 15% (95% CrI: 3%,7%) in 2012 and 18% (95% CrI: 7%, 70%) in 2013, interpreted as the annual incidence of DENV infection among children who were seronegative at the start of the year. These estimates suggest that during outbreak years, 15%–18% — or about 1 in 6 — children without evidence of prior DENV infection (seronegative at the start of the year) could be infected. Stratified analysis by health district revealed substantially higher transmission in St. Thomas/St. John (FOI in 2012–13: 36%–38%) compared to St. Croix (FOI in 2012–13: 7%–12%). In contrast, during non-outbreak years (2010–2011 and 2017–2022), estimated annual FOI was low (<1%), reflecting minimal ongoing transmission. The estimated reporting fraction—that is, the proportion of all DENV infections (symptomatic and asymptomatic) that resulted in a dengue disease case reported to ArboNET—was lower in St. Thomas/St. John (reporting fraction in 2012: 1.61%, 95% CrI: 0.69%, 4.47%) compared to St. Croix (reporting fraction in 2013: 10.0%, 95% CrI: 4.74%, 21.1%). An estimated 12% of reported cases were primary infections (95% CrI: 6%, 42%). For children born outside of USVI who previously lived in dengue-endemic countries, the annual FOI for the endemic countries was 6% (95% CrI: 3%, 10%), compared to 0% (95% CrI: 0%, 2%) for those previously residing in non-endemic areas, such as the continental U.S. Dengue-endemic countries or territories of prior residence included the Philippines, Dominica, Dominican Republic, Haiti, Anguilla, St. Lucia, Bermuda, and Puerto Rico.

**Figure 2.**
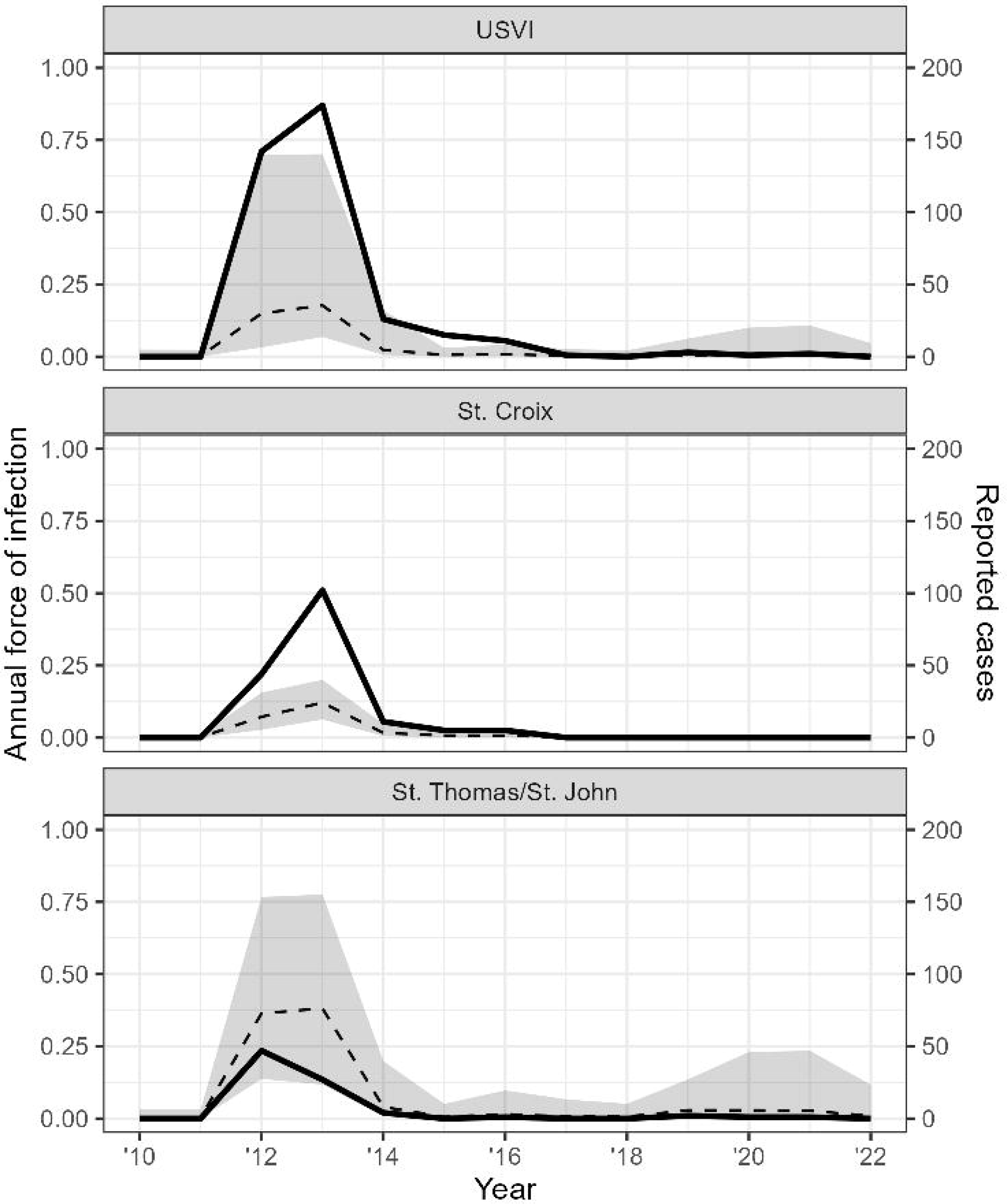
Estimated annual dengue force of infection for any serotype (dashed lines), and total cases reported (black lines), by health district and year — US Virgin Islands, 2010–2022. Estimates are derived from catalytic modeling using serosurvey data and age-stratified case reports. Transparent ribbons represent 95% CrI for annual FOI estimates.

Catalytic regression modeling revealed differences in the hazard of DENV infection by several participant characteristics (Supplemental Table C). Higher dengue hazard was observed among girls compared to boys (HR: 1.61, 95% CrI: 1.16, 2.32), among Asian/Pacific islander children compared to Black/African American children (HR: 6.66, 95% CrI: 1.97, 37.89), and Hispanic children compared to non-Hispanic children (HR 2.11, 95% CrI: 1.42, 3.26). Children attending private schools had a lower estimated hazard of DENV infection than those attending public schools, but this association was not statistically significant (HR 0.69, 95% CrI: 0.43, 1.07).

### Dengue vaccine knowledge, attitudes, and perceptions: univariate analysis

In April–May 2022, a total of 355 parents completed survey questions about dengue knowledge and attitudes toward dengue vaccination (Figure 3, Supplemental Table D). Overall, 82% (95% CI: 77%, 85%) of parents reported having heard of dengue. When asked whether their child had previously had dengue, most parents answered “no” (84%, 95% CI: 80%, 88%), and an additional 11% (95% CI: 8%, 15%) were unsure. Seventeen percent (95% CI: 14%, 22%) of parents were aware of an approved dengue vaccine recommended for children in the U.S. When asked about their willingness to vaccinate their child if a dengue vaccine became available in USVI, 25% (95% CI: 21%, 30%) indicated they would, 48% (95% CI: 42%, 53%) were unsure, and 27% (95% CI: 23%, 32%) reported they would not vaccinate their child.

**Figure 3.**
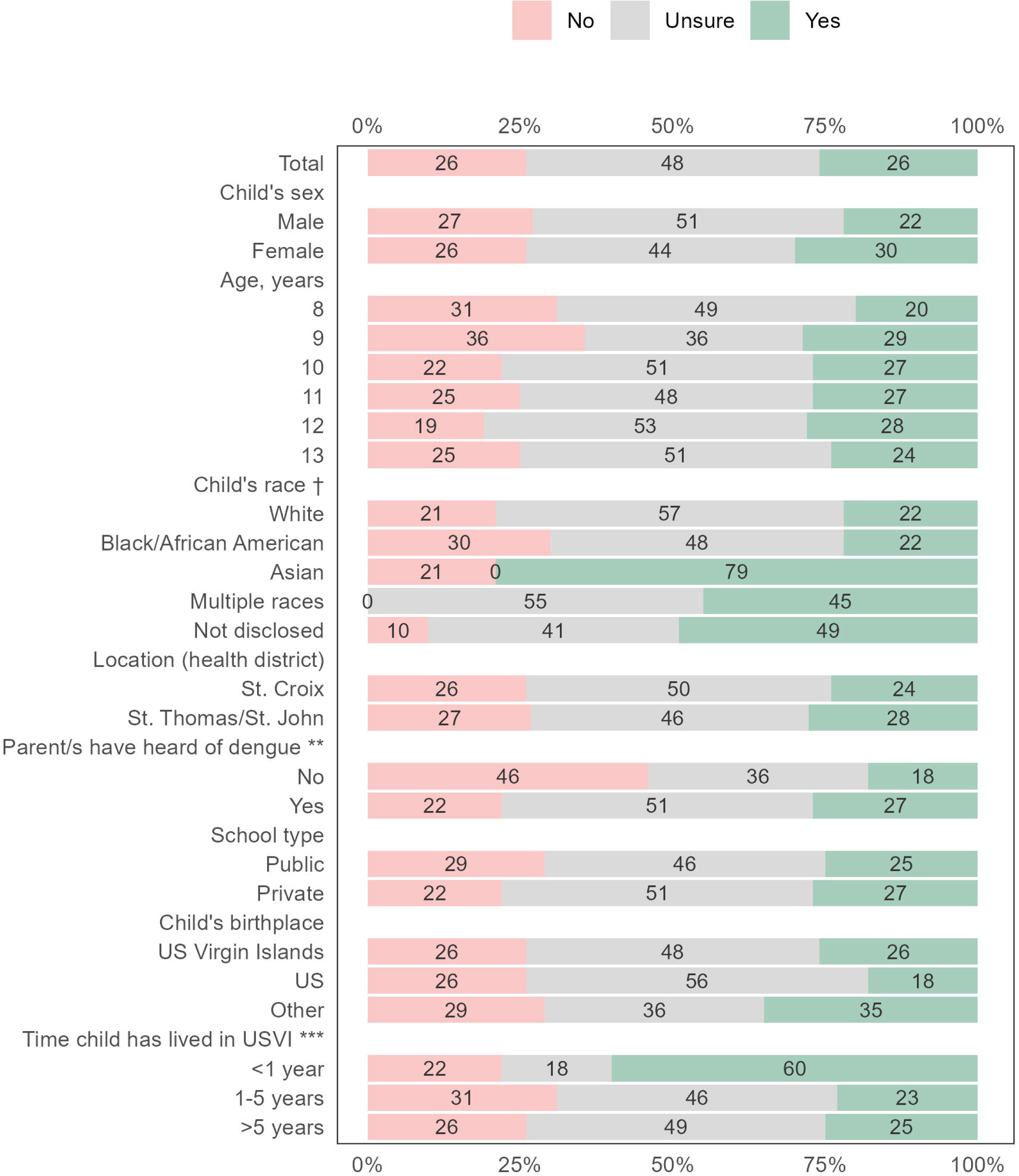
**Parental intention to vaccinate their child against dengue among serosurvey participants (N = 355*) — US Virgin Islands, April–May 2022.** *17 responses not included due to non-response to relevant survey questions ** Parents have heard of dengue: The distribution of responses (No/Unsure/Yes) differed significantly between parents who had heard of dengue and those who had not (22% vs 46% would not vaccinate) *** Time child has lived in USVI: Response distributions differed significantly for parents of children living in USVI <1 year versus longer durations. Estimates with corresponding 95% confidence intervals can be found in Supplemental Table E.

Parents who had heard of dengue were more likely than parents who had not heard of dengue to intend to vaccinate their children (27% vs 18%) or be undecided about vaccination (51% vs 36%) (Supplemental Table E). The ratio of parents that would not vaccinate to those that were unsure was statistically different between parents that had heard of dengue and those that had not (OR 0.33, 95% CI: 0.23, 0.49).

Parents whose children were born outside of the U.S. or USVI were more likely to intend to vaccinate (35%, 95% CI: 12%, 58%) compared to parents of children born in the mainland U.S. (26%, 95% CI: 19%, 33%) or born in the USVI (18%, 95% CI: 8%, 29%). Vaccine acceptance also varied by race: 79% (95% CI: 74%, 85%) of parents of Asian children would vaccinate, compared to only 22% (95% CI: 18%, 27%) of parents of Black children and 22% (95% CI: 5%, 39%) of parents of White children. No associated odds ratios were statistically significant for either birthplace or race. Parents of children that had lived in USVI for less than 1 year were more likely to reporting intending to vaccinate for dengue (60%, 95% CI: 35%, 85%) compared to parents of children that had lived in USVI for 1–5 years (23%, 95% CI: 6%, 40%) and for greater than 5 years (25%, 95% CI: 18%, 31%). This difference was statistically significant (1–5 years: OR 0.15, 95% CI: 0.02, 0.98; >5 years: OR 0.15, 95% CI: 0.03, 0.72). Full results can be found in Supplemental Table E.

## DISCUSSION

This study provides the first representative dengue seroprevalence estimates among school-aged children in both the St. Croix and St. Thomas/St. John health districts of USVI. Nearly half of participating children showed evidence of past DENV infection, despite most parents reporting that their child had never been diagnosed with dengue. This discrepancy may reflect underrecognition of symptomatic dengue illness as well as the high frequency of asymptomatic or mild infections that typically go undetected or unreported. These findings highlight the need for increased public awareness, improved prevention efforts, and strengthened laboratory capacity and case reporting for dengue as a nationally notifiable disease. The observed seroprevalence is valuable for modeling the potential impact of future dengue vaccination strategies in the territory. Low parental awareness of Dengvaxia, the only licensed and recommended dengue vaccine at the time, coupled with the substantial proportion of parents who were undecided about vaccinating their child if a dengue vaccine were available, suggests that investments in public education and community engagement will be crucial for successful implementation of future dengue vaccines.

In examining factors associated with previous DENV infection, we observed higher estimated seroprevalence in the St. Thomas/St. John health district (59%) compared to St. Croix (34%), although this difference was not statistically significant. The observed variation may reflect unmeasured differences in local *Aedes aegypti* ecology (e.g., abundance of water-holding containers, microclimate), housing density, and patterns of human mobility between districts, which were not directly captured in our data.^30^ After adjusting for health district and other covariates, children attending private schools had a lower estimated hazard of DENV infection than those attending public schools, although this association was not statistically significant. In the absence of direct measures of socioeconomic status (SES), school type may serve as a proxy, with private school enrollment potentially reflecting higher household income. However, other unmeasured factors—such as geographic clustering of schools, local mosquito abundance, and the highly overdispersed nature of dengue transmission—may also contribute to this association. During the 2016–2017 Zika outbreak in USVI, a study of pregnant women found low reported use of mosquito control measures such as bed nets and window screens.^31^ Although the role of cost was not directly assessed, approximately half of USVI residents live in rental housing, where home modifications may be out of residents’ control.^32^ Further research into SES-related risk factors in USVI could help guide equitable access to protective interventions and reduce disparities in arboviral disease risk.

The highest force of infection occurred during the well-documented dengue outbreak years of 2012 and 2013. During this period, we estimated a low probability of case reporting on St. Thomas/St. John (1.6%), similar to findings from Kada et al. in the U.S. and associated territories.^25^ This finding highlights the potential for substantial underdetection and underreporting of dengue in the USVI, consistent with the broader understanding that routine surveillance captures only a fraction of infections—particularly when many infections are asymptomatic or mild. Further investigation is needed into barriers to diagnosis and reporting—such as limited access to primary care, visit costs, transportation challenges, and time constraints for caregivers. Laboratory-related barriers, including limited testing availability, costs, and provider awareness or clinical suspicion of dengue, may also contribute to underdiagnosis and underreporting. Enhancing provider education, integrating syndromic surveillance, and improving laboratory capacity could help capture low-level transmission and detect periodic outbreaks that might otherwise go unrecognized.^5,33^ Given that many DENV infections are asymptomatic or mild, it is unsurprising that most parents reported their child had never had dengue or were unsure, despite serologic evidence of past infection in nearly half of participants. These findings support maintaining dengue prevention messaging, including mosquito bite avoidance, clinical vigilance, and strengthening on-island diagnostic capabilities, even during inter-epidemic periods.

The majority of parents reported awareness of dengue and indicated openness or uncertainty regarding vaccination. Implementation of a future dengue vaccine would likely require extensive outreach and educational campaigns to parents to provide them with the information needed to support dengue vaccination. The high seroprevalence findings indicate that school-age children are at high risk for secondary DENV infections, and prioritizing vaccination for this population – and the associated communication campaigns to parents and children – would be critical to reduce dengue burden. In our survey, parents who had previously heard of dengue and those with shorter residency in USVI demonstrated higher vaccine acceptance, suggesting that parents without prior awareness of dengue and long-term USVI residents may be more hesitant and thus require more intensive outreach and tailored messaging. Future research on health beliefs, motivations, and information needs about dengue vaccination within these subpopulations could provide valuable insights to local health officials, particularly as new dengue vaccines undergo licensure.

## Limitations

Limitations of this work include the inability to distinguish between single and multiple previous DENV infections based solely on IgG antibody testing, thus restricting a complete understanding of dengue burden in USVI children. The IgG test used also has imperfect sensitivity and specificity, so some misclassification of infection status is likely and could bias seroprevalence and FOI estimates, even though we incorporated published test performance characteristics into our estimation procedures. Additionally, the survey’s lack of open-ended questions about parental perceptions of dengue and dengue vaccines constrained deeper insights into parental attitudes and specific information needs.

## Public health implications

The high seroprevalence of dengue identified among school-aged children, despite low parental awareness of prior DENV infections, highlights an urgent need for improved public awareness and education about dengue in the USVI. The association between parental dengue awareness and vaccine acceptance emphasizes that targeted educational initiatives could significantly improve community engagement and acceptance of future dengue vaccination programs. These seroprevalence findings can directly inform public health planning, guiding the development of tailored educational campaigns, optimizing resource allocation, and supporting proactive dengue prevention strategies.

## Data Availability

Data are available from the CDC management team (contact) for researchers who meet the criteria for access to confidential data.

