## Supplement for "Sustained dengue transmission and seroprevalence in the U.S. Virgin Islands"

**Supplemental Material**

**Supplemental Methods.** Catalytic Model and Force of Infection Estimation

**Supplemental Table A.** Weighted seroprevalence of anti-DENV IgG antibodies by characteristic — US Virgin Islands, April–May 2022 (N=372)

**Supplemental Table B.** Annual confirmed dengue cases in USVI and estimated annual force of infection for all serotypes, 2010–2022. FOI estimates were derived from a catalytic model fit to confirmed case and serosurvey results

**Supplemental Table C**. Adjusted hazard ratios (HRs) for participant characteristics associated with dengue virus (DENV) infection, based on catalytic model regression — U.S. Virgin Islands, April–May 2022

**Supplemental Table D.** Parent-reported knowledge, perceptions, and vaccine acceptance related to dengue — U.S. Virgin Islands, April–May 2022

**Supplemental Table E.** Parental intention to vaccinate their child against dengue among serosurvey participants (N = 355) – US Virgin Islands, April–May 2022

**Supplemental Figure A.** Observed and catalytic model–estimated dengue seroprevalence and reported case counts by island, year, and age group — U.S. Virgin Islands

**Supplemental Methods: Catalytic Model and Force of Infection Estimation**

We excluded seven individuals from the catalytic model analysis due to unclear residence history in endemic versus non-endemic countries, resulting in a final sample of 365 participants. Models to estimate the annual dengue force of infection (FOI) were similar to those described by Adams et al. in their study of children in Puerto Rico.^1^ Briefly, we assumed that an individual’s seropositivity follows a Bernoulli distribution, with the probability of seropositivity determined by the cumulative hazard of DENV infection over their lifetime. Specifically, we defined the annual hazard of DENV infection for an individual residing in location *s* in year *t* as $\lambda_{t}^{s}$ , where *s* is one of *C, J, N,* or *E* (St. Croix, St. Thomas/St. John, another non-endemic country, or another endemic country). At the time of the serosurvey (*t=Y*), an individual *i* of age *a* and residence history $r_{t}^{i}$ experienced cumulative dengue FOI, calculated as: $\Lambda_{i}^{0}=\sum_{t=Y-a+1}^{Y} \lambda_{t}^{r_{t}^{i}}$. $r_{t}^{i}$ is a vector that represents an individual’s lifetime residence history; it has length *a* and each entry takes the value *C, J, N,* or *E* as above. We then allowed the cumulative hazard to vary by *R* individual-level covariates $x_{ir}$, assuming a multiplicative effect on the baseline hazard, following a log-linear model: $\log\left( \Lambda_{i} \right)=log(\Lambda_{i}^{0})+\sum_{r=1}^{R} \beta_{r}x_{ir}$. We assumed that hazard rates for non-endemic and endemic countries were constant over time and across countries. Additionally, we assumed that an individual’s island of residence within USVI remained constant throughout their lifetime, as the questionnaire did not capture inter-island migration.

To account for imperfect sensitivity and specificity of the diagnostic test, we modeled observed seropositivity as a function of both true positives (seropositive individuals who test positive) and true negatives (seronegative individuals who test negative).^2^ Sensitivity and specificity estimates were obtained from internal validations at CDC Dengue Branch using well-characterized specimens (CDC unpublished data). To account for the complex survey design, we assumed that all individuals were independent and that the sample was representative with respect to characteristics not already included in the survey weights. Model fitting was performed using a likelihood function weighted by the final design weights.^3^

Finally, we combined individual-level seropositivity data with age-stratified dengue case data from USVI to jointly estimate FOI and the reporting rate. Following the approach of Kada et al.,^4^ we derived the likelihood for the annual number of RT-PCR-confirmed dengue cases reported in each age group from 2010 to 2019 using ArboNET data, supplemented by local case counts from 2020–2022. For the years covered by the serosurvey, we estimated the island-specific FOI as a survey-weighted mean FOI using serosurvey participants residing on each island to account for sample non-representativeness. This island-wide FOI was then used to calculate the expected number of cases in each age group for each island group, assuming that cases represent a mixture of primary and secondary infections. We estimated annual reporting rate, meaning the proportion of all infections that are reported to ArboNET, and the relative reporting rate for primary vs. secondary infections, while interpreting FOI as the annual hazard of a first DENV infection among previously uninfected individuals. We jointly maximized the sum of the weighted log-likelihood from the serosurvey and the log-likelihood from the age-stratified case data.

Following Kada et al.,^4^ we partition variation in reporting rate and annual FOI into uncertainty in the mean and in year-to-year variability. We used a normal prior for the mean reporting rate with median 10% and 97.5^th^ percentile 30%, or mean -2.2 and standard deviation 0.49). We follow a process as outlined in Kada et al.^4^ to use data to inform the prior distribution of average inverse-logit FOI. For St. Croix, we used a normal prior with mean -4.8 and standard deviation 1.23, and for St. Thomas/St. John we used a normal prior with mean -5.8 and standard deviation 1.59. The relative reporting rate for primary vs. secondary infections had a uniform prior on [0,1], assuming that primary infections were less likely than secondary infections to be reported. For all regression coefficients we used a weakly informative uniform prior on [-4,4], while for the endemic and non-endemic annual FOI $\lambda_{t}^{N}$ and $\lambda_{t}^{E}$ we used a uniform prior on [-10,-0.7]. Finally, for the sensitivity and specificity of the serological test we used a uniform prior on [0.5,1]. Parameter estimation was performed using Bayesian Markov chain Monte Carlo (MCMC) methods implemented in the rstan package.^5^ We report median parameter estimates and associated 95% credible intervals from the posterior distributions. Convergence was assessed using the Gelman–Rubin diagnostic.^6^ All analyses were performed in R version 4.2.2.^6^

**Supplemental Table A.** Weighted seroprevalence^a^ of anti-DENV IgG antibodies by characteristic — US Virgin Islands, April–May 2022 (N=372).

|  | **US Virgin Islands** | **St. Croix** | **St. Thomas/St. John** |
| --- | --- | --- | --- |
| Characteristic | **Weighted Seroprevalence % (95% CI)** | **Weighted Seroprevalence  (%, 95% CI)** | **Weighted Seroprevalence  (%, 95% CI)** |
| **Overall, 8-13 year-olds (all survey participants)** | 47 (29, 68) | 34 (21, 50) | 59 (30, 86) |
| **Overall, 9-13 year-olds^b^ (Dengvaxia-eligible age group in this sample)** | 51 (39, 64) | 37 (22, 54) | 64 (40, 85) |
| **Age, years** | | | |
| 8 | 27 (17, 39) | 22 (1, 43) | 32 (18, 45) |
| 9 | 41 (16, 71) | 27 (15, 40) | 53 (16, 90) |
| 10 | 42 (26, 60) | 35 (14, 56) | 48 (28, 68) |
| 11 | 50 (24, 77) | 23 (0, 48) | 74 (21, 100) |
| 12 | 69 (45, 88) | 55 (25, 84) | 82 (66, 97) |
| 13 | 54 (18, 89) | 43 (1, 85) | 63 (0, 100) |
| **Sex** | | | |
| Female | 50 (22, 80) | 33 (17, 48) | 66 (32, 100) |
| Male | 45 (31, 59) | 36 (16, 56) | 52 (34, 70) |
| **Race** | | | |
| White | 33 (12, 53) | 25 (0, 61) | 41 (0, 100) |
| Black/African American | 49 (38, 60) | 40 (25, 54) | 57 (38, 76) |
| Asian/Pacific Islander | 69 (0, 100) | 0 (NA) | 100 (NA) |
| American Indian/ Alaska Native | NA (NA, NA) | 17 (0, 100) | 68 (NA, NA) |
| Multiple races | 38 (0, 100) | 17 (0, 100) | 68 (NA, NA) |
| Not disclosed | 47 (15, 79) | 12 (0, 37) | 94 (69, 100) |
| **Ethnicity** | | | |
| Hispanic or Latino | 58 (40, 76) | 44 (12, 75) | 79 (57, 1) |
| Non-Hispanic or Latino | 50 (35, 64) | 33 (19, 46) | 62 (37, 87) |
| **School type** | | | |
| Private | 33 (11, 55) | 28 (0, 63) | 45 (0, 100) |
| Public | 54 (40, 69) | 40 (18, 63) | 62 (41, 83) |
| **Time living in USVI** | | | |
| <1 year | 43 (0, 89) | 0 (0, 0) | 97 (0, 100) |
| 1-5 years | 36 (20, 51) | 5 (0, 22) | 58 (42, 73) |
| >5 years | 50 (37, 62) | 40 (23, 56) | 59 (37, 81) |
| **Birthplace** | | | |
| US | 34 (13, 54) | 25 (0, 58) | 46 (13, 8) |
| USVI | 50 (39, 61) | 39 (25, 54) | 60 (41, 8) |
| Other | 50 (32, 69) | 2 (0, 16) | 63 (41, 84) |

Abbreviations: CI=confidence interval, NA=unable to calculate

^a^ Percentage estimates are weighted and are standardized to the age and sex of the 2022 U.S. Census population estimated distribution across the two districts.

^b^ Children aged 8 years are not currently eligible for Dengvaxia under ACIP recommendations; this row reflects parental intention among the Dengvaxia-eligible age subset (9–13 years) within the serosurvey.

**Supplemental Table B.** Annual confirmed dengue cases in USVI and estimated annual force of infection for all serotypes, 2010–2022. FOI estimates were derived from a catalytic model fit to confirmed case and serosurvey results. Estimates are shown for: (1) all serosurvey participants, and (2) participants stratified by health district (St. Croix and St. Thomas/St. John).

| Year | Confirmed cases reported in USVI^a^ | FOI % (95% CrI),  USVI (all health districts combined) | FOI % (95% CrI),  St. Croix | FOI % (95% CrI),  St. Thomas/ St. John |
| --- | --- | --- | --- | --- |
| 2010 | 0 | 0 (0,2) | 0 (0,1) | 0 (0,3) |
| 2011 | 0 | 0 (0,2) | 0 (0,1) | 0 (0,3) |
| 2012 | 142 | 15 (3,70) | 7 (3,16) | 36 (14,77) |
| 2013 | 174 | 18 (7,70) | 12 (6,20) | 38 (12,78) |
| 2014 | 26 | 2 (1,16) | 2 (1,5) | 4 (1,20) |
| 2015 | 15 | 1 (0,3) | 1 (0,3) | 0 (0,3) |
| 2016 | 11 | 1 (0,4) | 1 (0,3) | 1(0,5) |
| 2017 | <3 | 0 (0,3) | 0 (0,1) | 0 (0,3) |
| 2018 | 0 | 0 (0,2) | 0 (0,1) | 0 (0,3) |
| 2019 | 3 | 1 (0,6) | 0 (0,1) | 2 (0,8) |
| 2020 | <3 | 0 (0,10) | 0 (0,1) | 2 (0,13) |
| 2021 | <3 | 0 (0,11) | 0 (0,1) | 2 (0,13) |
| 2022 | 0 | 0 (0,5) | 0 (0,1) | 1 (0,7) |
| Country of Previous Residence (non-USVI) | | | | |
| Endemic^b^ | 6 (3,10) | --- | --- | --- |

**Acronyms**: CrI = Credible interval, FOI = Force of infection, USVI = US Virgin Islands

FOI estimates were generated from a catalytic model jointly fit to confirmed dengue case and serosurvey results. For the full analysis, 365 children were included; 7 were excluded due to missing residence history. The lifelong resident analysis included 266 children whose parents indicated they were born in the USVI; 99 children born elsewhere were excluded.

^a^ Confirmed cases were obtained from ArboNET and local surveillance data and classified according to the 2015 CSTE dengue case definition.

^b^ Among children born elsewhere, FOI was substantially higher for those from dengue-endemic countries and nearly zero for those from non-endemic regions, including the continental U.S.

**Supplemental Table C.** Adjusted hazard ratios (HRs) for participant characteristics associated with dengue virus (DENV) infection, based on catalytic model regression — U.S. Virgin Islands, April–May 2022.

|  | **Adjusted HR (95% CI)** |
| --- | --- |
| **Sex^a^** |  |
| Male | Ref |
| Female | 1.61 (1.16,2.32) |
| **Race^a^** |  |
| Black/African American | Ref |
| White | 0.74 (0.29,1.56) |
| Asian/Pacific Islander | 6.66 (1.97,37.89) |
| Multiple races | 0.97 (0.27,3.29) |
| Other or missing | 1.02 (0.54,1.93) |
| **Ethnicity^a^** |  |
| Non-Hispanic or Latino/ Other/missing | Ref |
| Hispanic | 2.11 (1.42,3.26) |
| **School Type^a^** |  |
| Public | Ref |
| Private | 0.69 (0.43,1.07) |

**^a^** Adjusted for Health District (i.e., St. Croix or St. Thomas / St. John), non-USVI residency, and all other covariates

**Supplemental Table D.** Parent-reported knowledge, perceptions, and vaccine acceptance related to dengue — U.S. Virgin Islands, April–May 2022.

|  | **Total Sample**  **N (%)** | **St. Croix**  **n (%)** | **St. Thomas/ St. John**  **n (%)** |
| --- | --- | --- | --- |
| **Total** | 355 (100)^a^ | 188 (100) | 167 (100) |
| **Response to Question**  **“Have you ever heard about dengue before?”** (N=354) | | | |
| Yes | 289 (82) | 166 (88) | 123 (74) |
| No | 65 (18) | 22 (12) | 43 (26) |
| **Response to Question**  **“Has your child ever had dengue?”** | | | |
| Yes | 3 (1) | 3 (2) | 0 (0) |
| No | 299 (84) | 159 (85) | 140 (84) |
| I don’t know | 38 (11) | 22 (12) | 16 (10) |
| I’ve never heard about dengue before | 15 (4) | 4 (2) | 11 (7) |
| **Response to Question**  **“Did you know there is an approved dengue vaccine that was recommended for children in the US?”** (N=354) | | | |
| Yes | 61 (17) | 34 (18) | 27 (16) |
| No | 293 (83) | 153 (82) | 140 (84) |
| **Response to Question**  **“If this approved dengue vaccine for children was made available in USVI, would you have your child vaccinated?”** | | | |
| Yes | 90 (25) | 42 (22) | 48 (29) |
| No | 96 (27) | 51 (27) | 45 (27) |
| Undecided | 169 (48) | 95 (51) | 74 (44) |

^a^ Total sample size was 355. Some questions had one missing or incomplete response, as indicated by (N=354).

**Supplemental Table E:** Parental intention to vaccinate their child against dengue among serosurvey participants (N = 355) – US Virgin Islands, April–May 2022.

|  | **No intention to vaccinate** | **Undecided about vaccination** | **Do intend to vaccinate** |  |  |
| --- | --- | --- | --- | --- | --- |
| Characteristic | **Weighted %**  **(95% CI)** | **Weighted %**  **(95% CI)** | **Weighted %**  **(95% CI)** | **OR (95% CI) (No:Undecided vs reference group)** | **OR (95% CI) (Yes:Undecided vs reference group)** |
| **Overall, 8-13 year-olds (all survey participants)** | 26 (24, 29) | 48 (42, 54) | 26 (20, 31) |  |  |
| **Overall, 9-13 year-olds^a^ (Dengvaxia-eligible age group in this sample)** | 25 (22, 28) | 48 (42, 54) | 27 (20, 33) |  |  |
| **Age, years** | | | |  |  |
| 8 | 31 (19, 44) | 49 (38, 60) | 20 (10, 30) | REF | REF |
| 9 | 36 (28, 43) | 36 (21, 50) | 29 (20, 38) | 0.63 (0.26, 1.57) | 0.51 (0.18, 1.39) |
| 10 | 22 (16, 29) | 51 (43, 60) | 27 (18, 35) | 1.43 (0.68, 3.06) | 0.79 (0.34, 1.84) |
| 11 | 25 (13, 38) | 48 (34, 61) | 27 (14, 40) | 1.19 (0.45, 3.18) | 0.73 (0.27, 2.03) |
| 12 | 19 (10, 29) | 53 (36, 69) | 28 (11, 45) | 1.70 (0.65, 4.45) | 0.77 (0.24, 2.52) |
| 13 | 25 (11, 39) | 51 (31, 71) | 24 (7, 41) | 1.29 (0.42, 4.02) | 0.87 (0.24, 3.10) |
| **Sex** | | | |  |  |
| Male | 27 (21, 33) | 51 (42, 60) | 22 (17, 26) | REF | REF |
| Female | 26 (21, 31) | 44 (36, 53) | 30 (21, 39) | 0.91 (0.54, 1.56) | 0.63 (0.32, 1.21) |
| **Child’s race^b^** | | | |  |  |
| White | 21 (4, 37) | 57 (46, 68) | 22 (5, 39) | --- | REF |
| Black/African American | 30 (27, 32) | 48 (43, 53) | 22 (18, 27) | --- | 0.92 (0.59, 1.43) |
| Asian/Pacific Islander | 21 (15, 26) | 0 (0, 0) | 79 (74, 85) | --- | 1.32 (0.87, 2.01) |
| Multiple races | 0 (0, 0) | 55 (21, 89) | 45 (11, 79) | --- | 1.62 (1.06, 2.48) |
| Not disclosed | 10 (0, 20) | 41 (17, 64) | 49 (21, 77) | --- | 1.37 (0.79, 2.38 |
| **Health district** | | | | | |
| St. Croix | 26 (23, 30) | 50 (40, 60) | 24 (14, 33) | REF | REF |
| St Thomas/St. John | 27 (23, 31) | 46 (40, 52) | 27 (21, 32) | 1.09 (0.74, 1.67) | 1.30 (0.63, 2.72) |
| **Parent has heard of dengue** | | | |  |  |
| No | 46 (39, 53) | 36 (29, 43) | 18 (9, 27) | REF | REF |
| Yes | 22 (18, 26) | 51 (44, 59) | 27 (19, 35) | **0.33 (0.23, 0.49)** | 1.00 (0.45, 2.31) |
| **School type** | | | |  |  |
| Public | 29 (25, 32) | 46 (41, 51) | 25 (21, 30) | REF | REF |
| Private | 22 (17, 27) | 51 (39, 63) | 27 (17, 37) | 0.71 (0.43, 1.29) | 0.95 (0.46, 1.95) |
| **Child’s birthplace** | | | |  |  |
| US Virgin Islands | 26 (14, 37) | 56 (41, 71) | 18 (7, 30) | REF | REF |
| United States | 26 (21, 31) | 48 (41, 55) | 26 (19, 33) | 1.17 (0.54, 2.62) | 1.64 (0.60, 4.29) |
| Other | 29 (15, 44) | 36 (23, 49) | 35 (12, 59) | 1.80 (0.75, 4.37) | 3.00 (0.73, 12.09) |
| **Time child has lived in USVI** | | | |  |  |
| <1 year | 22 (1, 42) | 18 (0, 38) | 60 (35, 85) | REF | REF |
| 1-5 years | 31 (20, 42) | 46 (31, 62) | 23 (6, 40) | 0.55 (0.09, 3.61) | **0.15 (0.02, 0.98)** |
| >5 years | 26 (22, 29) | 49 (43, 56) | 25 (18, 31) | 0.44 (0.07, 2.64) | **0.15 (0.03, 0.72)** |

^a^ Children aged 8 years are not currently eligible for Dengvaxia under ACIP recommendations; this row summarizes parental intention among the age-eligible subset (9–13 years) within the serosurvey.
^b^ Odds ratios comparing Yes:No responses were estimated for race categories.

**Supplemental Figure A.** Observed and catalytic model–estimated dengue seroprevalence and reported case counts by island, year, and age group — U.S. Virgin Islands.

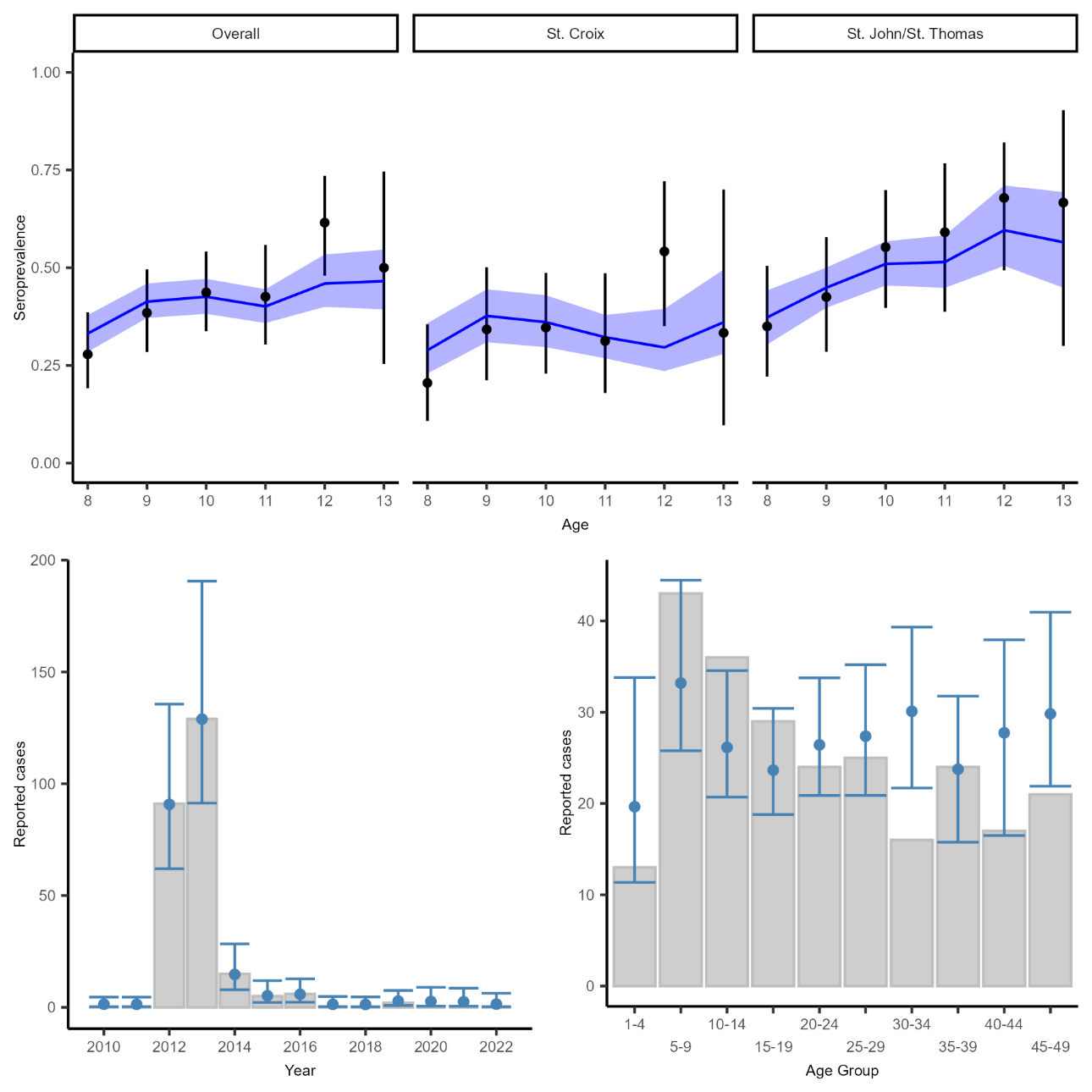

Top row: Estimated age-specific dengue seroprevalence by island among children aged 8–13 years, based on a catalytic model that infers force of infection from the current serosurvey and reported case data. Black points and lines represent observed age-specific seroprevalence and associated 95% CIs. Blue lines represent model-estimated, weighted and sensitivity/specificity-adjusted seroprevalence, with blue ribbons representing associated 95% CrI. Bottom row: Observed (gray bars) and estimated (blue points) annual dengue cases by year (left) and age group (right). Case data shown include all age groups (not limited to 8–13 years), as all reported case ages were used to inform the model. Blue error bars represent 95% credible intervals.
